# Compartment-specific Fat Distribution Profiles have Distinct Relationships with Cardiovascular Ageing and Future Cardiovascular Events

**DOI:** 10.1101/2025.09.16.25335879

**Authors:** Cynthia Maldonado-Garcia, Ahmed Salih, Stefan Neubauer, Steffen E. Petersen, Zahra Raisi-Estabragh

## Abstract

Obesity is a global public health priority and a major risk factor for cardiovascular disease (CVD). Emerging evidence indicates variation in pathologic consequences of obesity deposition across different body compartments. Biological heart age may be estimated from imaging measures of cardiac structure and function and captures risk beyond traditional measures. Using cardiac and abdominal magnetic resonance imaging (MRI) from 34,496 UK Biobank participants and linked health record data, we investigated how compartment-specific obesity phenotypes relate to cardiac ageing and incident CVD risk. Biological heart age was estimated using machine learning from 56 cardiac MRI phenotypes. K-means clustering of abdominal visceral (VAT), abdominal subcutaneous (ASAT), and pericardial (PAT) adiposity identified a high-risk cluster (characterised by greater adiposity across all three depots) associated with accelerated cardiac ageing – and a lower-risk cluster linked to decelerated ageing. These clusters provided more precise stratification of cardiovascular ageing trajectories than established body mass index categories. Mediation analysis showed that VAT and PAT explained 13.7% and 11.9% of obesity-associated CVD risk, respectively, whereas ASAT contributed minimally, with effects more pronounced in males. Thus, cardiovascular risk appears to be driven primarily by visceral and pericardial rather than subcutaneous fat. Our findings reveal a distinct risk profile of compartment-specific fat distributions and show the importance of pericardial and visceral fat as drivers of greater cardiovascular ageing. Advanced image-defined adiposity profiling may enhance CVD risk prediction beyond anthropometric measures and enhance mechanistic understanding.

## Introduction

Obesity is a global public health crisis. Projections indicate that by 2030, over 1 billion people will be affected by obesity [1]. While anthropometric measures such as, body mass index (BMI), are routinely used to quantify obesity, they fail to capture the complexity of adiposity distribution patterns and their metabolic consequences [2,3].

Advances in imaging technologies have revolutionised the assessment of adiposity by enabling precise, compartment-specific quantification of fat distribution (e.g., visceral, subcutaneous, and ectopic fat deposition) [4]. These techniques reveal that not all fat depositions are equal – with variations in distribution patterns strongly linked to disparate cardiometabolic outcomes [5,6]. The recent proliferation of large-scale imaging biobanks offers unprecedented opportunities to investigate obesity phenotypes and their longitudinal health implications.

Obesity is a well-established risk factor for cardiovascular disease (CVD) [7,8]. Preliminary studies suggest that distinct adiposity patterns correlate with varying degrees of cardiac dysfunction, vascular changes, and metabolic dysregulation [9,10]. Longitudinal analyses are not well explored, and underlying mechanisms are poorly understood.

Biological heart age reflects structural and functional changes in the cardiovascular system that occur with ageing, independent of chronological age [11,12]. This phenotype is quantified using imaging-derived measures of cardiac structure and function [13,14]. Emerging research suggests that cardiac ageing phenotypes may refine cardiovascular risk prediction, potentially identifying subclinical dysfunction not captured by conventional risk factors.

This study investigated the value of compartment-specific obesity phenotyping for refining risk stratification beyond anthropometric measures and examined variation in adverse consequences of different obesity depots by considering their relationships with biological heart ageing and incident clinical outcomes. The analysis integrates large-scale anthropometric and imaging-based adiposity profiling, cardiovascular magnetic resonance (CMR), and prospectively ascertained incident cardiovascular events – while considering variations in observed relationships across sexes.

## Results

### Baseline characteristics

From an initial pool of 44,930 UK Biobank participants with CMR data, we excluded 6,505 (14.5%) with >5% missing data and 3,929 (8.7%) with prevalent CVD, yielding a final analytical sample of 34,496 participants (18,978 females, 15,518 males) (**Figure S1**). The median age at imaging was 64 ± 7.9 years for males and 63.1 ± 7.6 years for females (mean ± SD). Most participants were from White (>96%) ethnic backgrounds.

Females had, on average, lower body mass index (BMI), waist circumference, and hip circumference than males. Over 66% of the males, had a BMI that was in the overweight BMI category or higher, while for females this figure was just over 50%. Females had a greater proportion of individuals at the extreme BMI categories (underweight and severe obesity) than males.

Females exhibit greater abdominal subcutaneous adipose tissue (ASAT), with volumes approximately 36% higher than those observed in males. In contrast, males possess markedly greater visceral adipose tissue (VAT) and pericardial adipose tissue (PAT), averaging 86% and 55% higher, respectively.

Across all BMI weight categories, VAT remains consistently ∼1.7-fold greater in males. In females, ASAT exceeds males ASAT by approximately 1.3–1.4-fold in the normal weight and overweight categories; however, this difference narrows in severe obesity. PAT volumes are consistently 1.4–1.5-fold greater in males, with the disparity most pronounced in the obesity category. Complete baseline characteristics for all study participants and a flow chart of study analysis steps are provided in **Table 1** and **Figure 1**.

**Figure 1.**
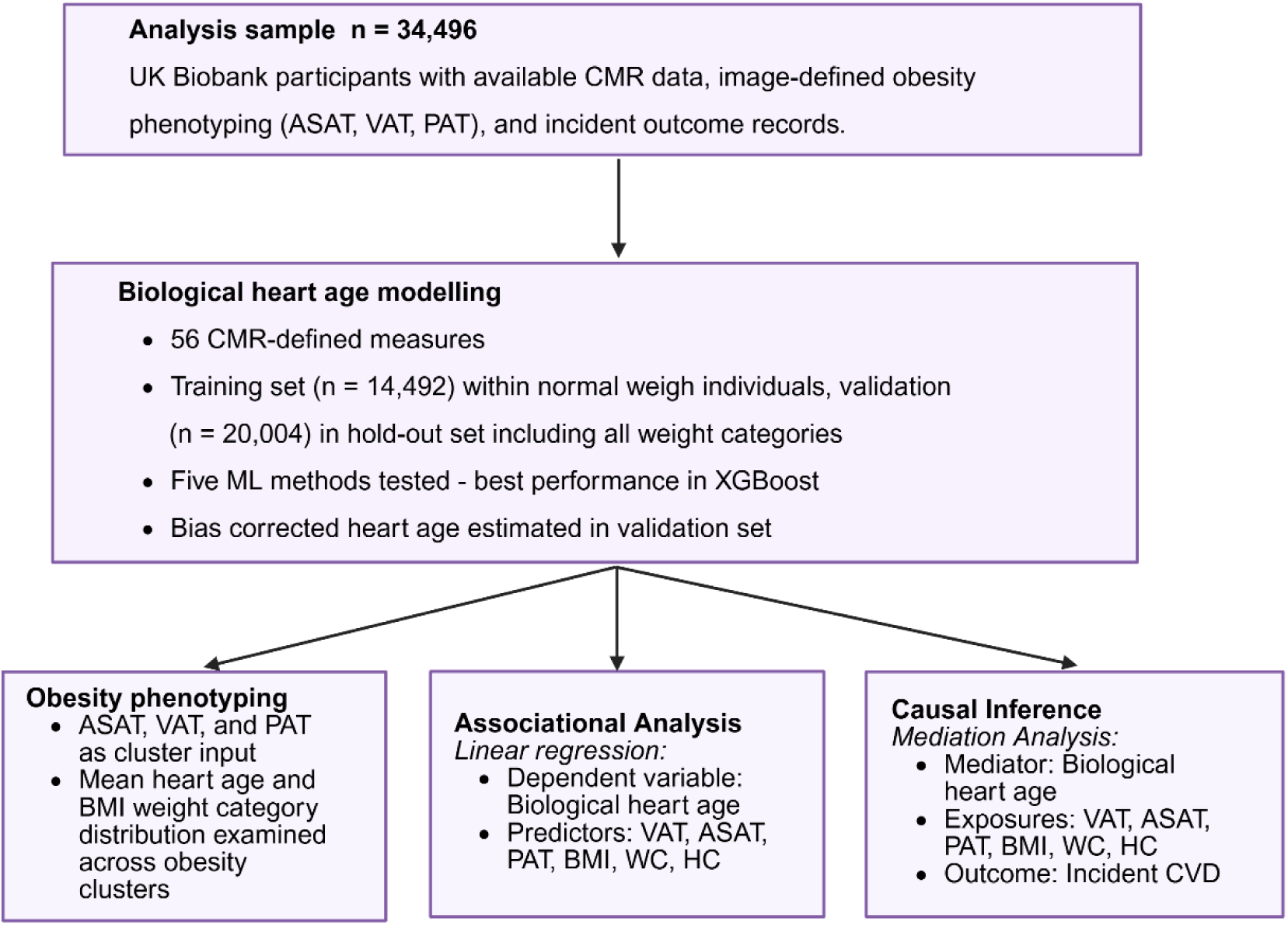
Flowchart illustrating the main steps for predicting biological heart age and the analyses performed. CMR, cardiac magnetic resonance; VAT, visceral adipose tissue; ASAT, abdominal subcutaneous adipose tissue; PAT, pericardial adipose tissue; BMI, body mass index; WC, waist circumference; HC, hip circumference; CVD, cardiovascular disease.

**Table 1.** Baseline characteristics of the study population stratified by sex. Values are presented as mean *±* standard deviation for continuous variables and percentages for categorical variables. Abbreviations: SBP, Systolic Blood Pressure; BMI, Body Mass Index; HC, Hip Circumference; WC: Waist Circumference; GCSEs, General Certificate of Secondary Education; CSEs, Certificate of Secondary Education; NVQ, National Vocational Qualification; HND, Higher National Diploma; HNC, Higher National Certificate; CVD: Cardiovascular Diseases.

|  | Male | Female |
| --- | --- | --- |
| n | 15,518 | 18,978 |
| Age (years) | 64.03 ± 7.87 | 63.09 ± 7.57 |
| Smoking status |  |  |
| Never (%) | 60.16 | 66.18 |
| Previous (%) | 35.45 | 30.71 |
| Current (%) | 4.17 | 2.76 |
| Prefer not to answer (%) | 0.21 | 0.35 |
| Number of days per week of moderate physical activity 10+ minutes (days) | 3.9 ± 2.25 | 3.99 ± 2.33 |
| BMI (kg/m<sup>2</sup>) | 26.86 ± 3.82 | 25.89 ± 4.69 |
| Townsend Deprivation index | -1.95 ± 2.71 | -1.82 ± 2.74 |
| SBP (mmHg) | 144.06 ± 18.48 | 138.03 ± 20.81 |
| WC (cm) | 93.62 ± 10.50 | 82.30 ± 11.72 |
| HC (cm) | 100.41 ± 7.26 | 100.59 ± 9.76 |
| Ethnicity |  |  |
| White (%) | 96.51 | 96.72 |
| South Asian (%) | 1.42 | 0.79 |
| Black (%) | 0.69 | 0.73 |
| Chinese (%) | 0.23 | 0.36 |
| Mixed (%) | 0.37 | 0.61 |
| Other (%) | 0.46 | 0.57 |
| Unknown (%) | 0.32 | 0.22 |
| Highest qualification category |  |  |
| College/university degree (%) | 53.26 | 51.15 |
| A levels/AS levels or equivalent (%) | 13.25 | 14.99 |
| O levels/GCSEs or equivalent (%) | 16.70 | 19.65 |
| CSEs or equivalent (%) | 3.08 | 2.63 |
| NVQ/HND/HNC or equivalent (%) | 4.68 | 1.78 |
| Other professional qualifications e.g.: nursing, teaching (%) | 1.41 | 2.29 |
| None of the above (%) | 7.24 | 7.13 |
| Prefer not to answer (%) | 0.40 | 0.38 |
| BMI obesity |  |  |
| Underweight (%) | 0.23 | 1.40 |
| Normal weight (%) | 33.14 | 46.86 |
| Overweight (%) | 47.58 | 33.61 |
| Obesity (%) | 16.54 | 15.44 |
| Severe Obesity (%) | 2.50 | 2.68 |
| Image-defined obesity compartments |  |  |
| Abdominal subcutaneous adipose tissue (L) | 5.68 ± 2.44 | 7.73 ± 3.34 |
| Visceral Adipose Tissue (mL) | 28.42 ± 13.97 | 18.28 ± 8.69 |
| Pericardial Adipose Tissue (cm<sup>2</sup>) | 4.73 ± 2.28 | 2.54 ± 1.48 |
| Number of incident CVD events | 811 | 550 |

### Biological Heart Age Prediction Modelling

The XGBoost model demonstrated superior performance compared to the other ML methods (**Table S1**). Our sex-specific XGBoost models, trained on normal weight participants (n=7,432 females; n=4,162 for males), demonstrated robust performance in holdout test sets (n=1,858 for females n=1,040 for males), achieving mean absolute errors of 4.94 years (R²=0.34) in females and 5.56 years (R²=0.27) in males. Post-bias-correction analysis confirmed successful age-adjustment, with near-zero correlations between heart age delta and chronological age (r=0.02 females, r=0.05 males) while maintaining strong associations between predicted and actual heart age (r=0.89 females, r=0.92 males), confirming age-independent detection of biological heart age (**Table S1**).

Sex-specific patterns emerged in CMR-derived feature importance in relation to biological age (**Figure 2**). Left ventricular mass (LVM) ranked highest in males and was among the most informative features in females. In females, left ventricular (LV) measures were most important in defining biological heart age, with LV end-diastolic volume (LVEDV) and end-systolic volume (LVESV) ranking among the most important predictors. In contrast, males showed greater reliance on right atrial (RA) parameters. Notably, six of the top ten predictive features in females reflected LV structure or function, compared to only three in males.

**Figure 2.**
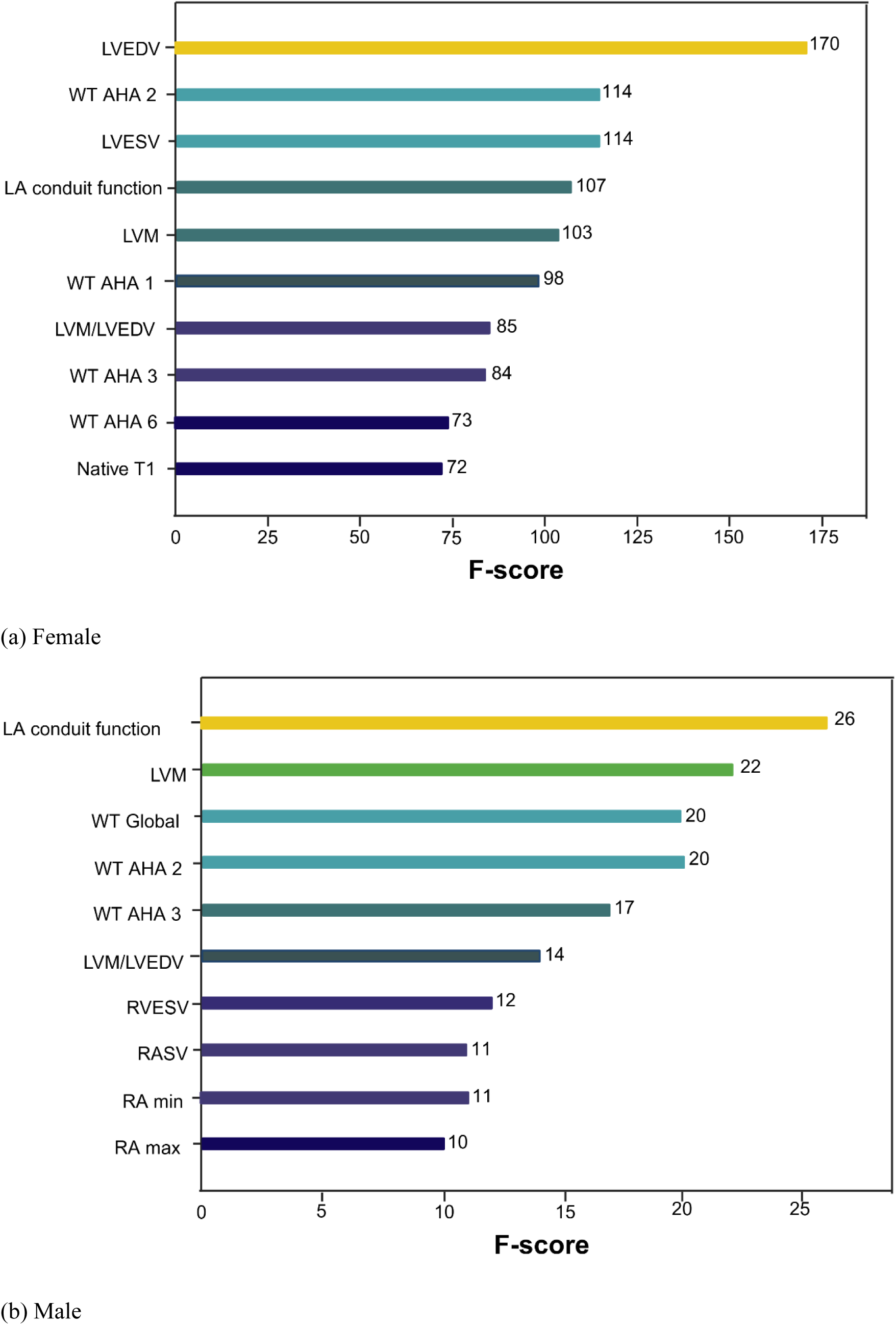
Top ten feature importances for predicting age using the XGBoost model, shown separately for females (a) and males (b). Features are ranked by their relative contribution to the model represented by the F-score. LVEDV, Left Ventricular End-Diastolic Volume; LVESV, Left Ventricular End-Systolic Volume; LVM, Left Ventricular Mass; LVM/LVEDV, the ratio of Left Ventricular Mass to Left Ventricular End-Diastolic Volume; WT AHA 1, Left Ventricular Wall Thickness 1; WT AHA 2, Left Ventricular Wall Thickness 2; WT AHA 3, Left Ventricular Wall Thickness 3; WT AHA 6, Left Ventricular Wall Thickness 6; WT Global, Left Ventricular Wall Thickness Global; LVGLS, Left Ventricular Global Longitudinal Strain; LA conduit function, Left Atrial Conduit Volume; RVESV, Right Ventricular End-Diastolic Volume; RASV, Right Atrial Stroke Volume; RA min, Right Atrial Minium Volume; RA max, Right Atrial Maximum Volume.

### Biological Heart Ageing Across Obesity Phenotypes

We identified distinct sex-specific patterns in heart age delta across BMI categories (**Figure 3**). Both sexes showed the most favourable cardiovascular ageing profiles in normal weight participants (mean heart age delta: -0.09 years in females, -0.03 years in males), consistent with healthier biological ageing. Females had a graded increase in heart age delta from normal weight to severe obesity (normal weight: -0.09 → underweight: 0.05 → overweight: 0.11 → obese: 0.22 → severe obesity: 0.58 years) (**Figure 3a**). Males displayed a non-linear pattern with peak heart age delta in the obese category (normal weight: -0.03 → underweight: 0.08 → overweight: 0.17 → severe obesity: 0.36 → obese: 0.60 years) (**Figure 3b**).

**Figure 3.**
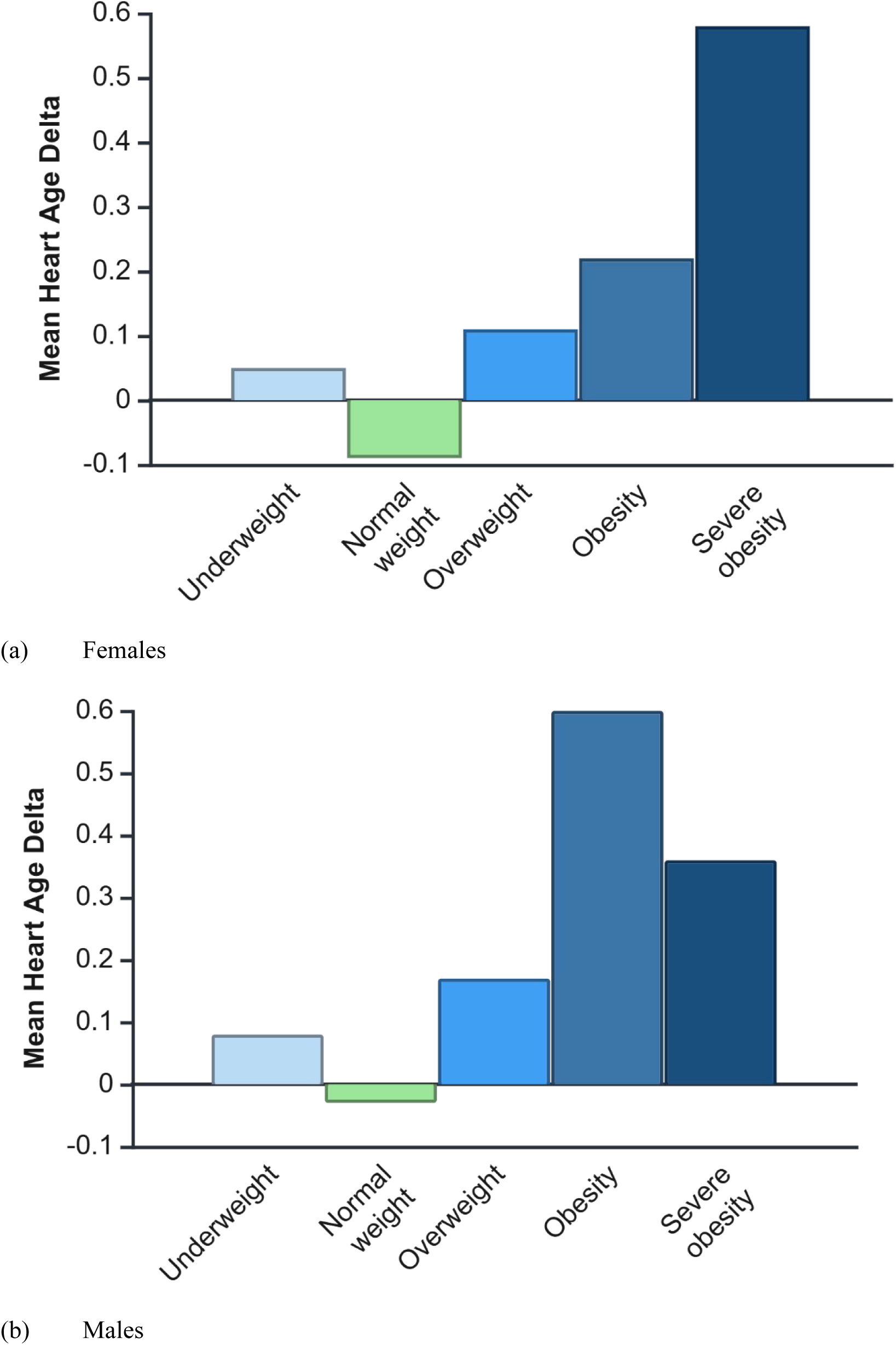
Sex-stratified comparison of heart age delta across BMI categories: underweight <18.5 kg/m2), normal weight (18.5–24.9 kg/m2), overweight (25–29.9 kg/m2), obese (30–39.9 kg/m2), and severely obese (≥40 kg/m2). (a) Mean heart age delta in females. (b) Mean heart age delta in males. Positive values indicate greater cardiac ageing relative to chronological age; negative values indicate less cardiac ageing.

Using the elbow method and silhouette analysis (**Figure S2**), we identified two distinct image-defined adiposity groups (**Figure 4a** and **Figure 4c**). The high-risk Cluster 0 was characterised by greater cardiovascular ageing, showing positive mean heart age delta values (+0.41 years in females, +0.71 years in males) and being composed of all severe obesity cases for each sex. This cluster exhibited significantly higher adipose tissue measures across all measured depots (**Table 2**). In contrast, the low-risk Cluster 1 demonstrated protective cardiovascular ageing patterns with negative mean heart age delta values (-0.15 years in females, -0.01 years in males) and substantially lower fat deposition across all fat compartments (**Table 2**). While most people fell into the cluster expected for their BMI group, there were some instances of mis-categorisation. For example, 11% of females and 9% of males in the normal BMI category were in the high-risk image-defined obesity cluster (**Figure 4**).

**Figure 4.**
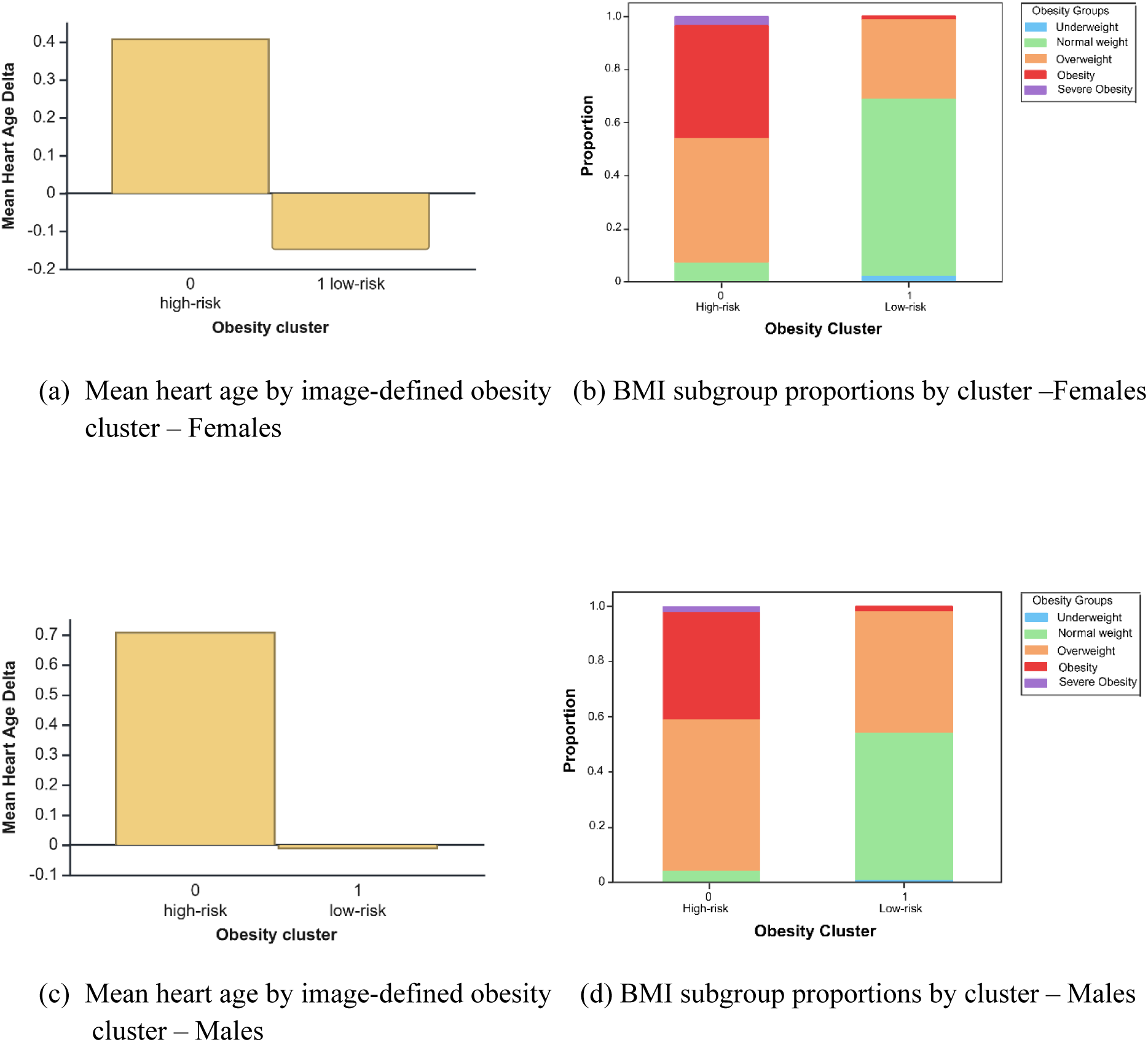
Mean heart age and BMI-subgroup distribution across phenotypic clusters, stratified by sex. Subfigures (a) and (c) show the mean heart age for Cluster 0 (high-risk) and Cluster 1 (protected) in females and males, respectively. Subfigures (b) and (d) show the proportion of BMI subgroups (underweight, normal weight, overweight, obesity, severe obesity) within each cluster for females and males, respectively.

**Table 2.**
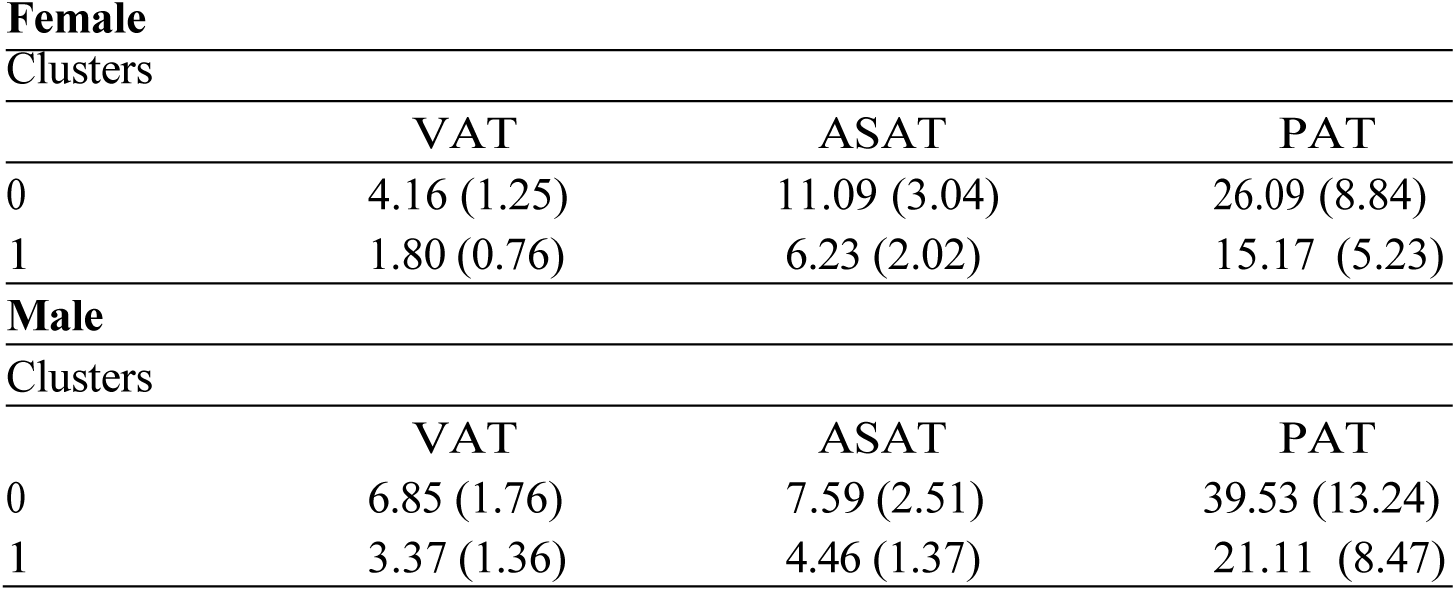
Sex-stratified adiposity measures (mean ± SD) across phenotypic clusters. Cluster 0: high-risk phenotype. Cluster 1: protected phenotype. Units: VAT (mL), ASAT (L), PAT (cm^2^). Abbreviations: VAT, visceral adipose tissue; ASAT, abdominal subcutaneous adipose tissue; PAT, pericardial adipose tissue.

Female
| Clusters | VAT | ASAT | PAT |
| --- | --- | --- | --- |
| 0 | 4.16 (1.25) | 11.09 (3.04) | 26.09 (8.84) |
| 1 | 1.80 (0.76) | 6.23 (2.02) | 15.17 (5.23) |

**Table 2. Sex-stratified adiposity measures (mean ± SD) across phenotypic clusters.**
| Clusters | VAT | ASAT | PAT |
| --- | --- | --- | --- |
| 0 | 6.85 (1.76) | 7.59 (2.51) | 39.53 (13.24) |
| 1 | 3.37 (1.36) | 4.46 (1.37) | 21.11 (8.47) |

Among female participants, high-risk Cluster 0 consistently demonstrated higher adipose tissue metrics compared to Cluster 1 within each BMI category. For obese females in Cluster 1, mean VAT was 2.49 ± 0.61 L versus 4.57 ± 1.28 L in Cluster 0, while ASAT showed similar differentials (9.91 ± 1.40 L vs 12.76 ± 2.36 L). The most striking contrast emerged in PAT, with Cluster 1 obese females averaging 14.20 ± 3.80 mL compared to 26.22 ± 9.73 mL in Cluster 0. This pattern held across all BMI categories, with normal weight Cluster 1 participants showing VAT levels 51% lower (1.60 ± 0.69 L) than their Cluster 0 counterparts (3.28 ± 0.78 L) (**Table S2**).

Male participants exhibited parallel but quantitatively distinct patterns. The low-risk Cluster 1 displayed substantially lower adiposity measures than Cluster 0 across all obesity categories. For instance, obese males in Cluster 1 had mean VAT of 4.86 ± 1.03 L compared to 7.64 ± 1.76 L in Cluster 0, with even more pronounced differences in PAT (21.32 ± 6.04 mL vs 40.15 ± 14.53 mL). Notably, normal weight males in Cluster 0 showed paradoxically elevated VAT (5.55 ± 1.10 L) compared to Cluster 1, suggesting potential limitations of BMI classification in males. There was a complete absence of severely obese participants in the low-risk clusters for both sexes (0 cases in Cluster 1) (**Table S2**).

### Associations with Biological Heart Age

Our analysis revealed distinct sex-specific patterns in the relationship between body fat distribution and greater heart ageing. All anthropometric and image-defined obesity measures showed statistically significant associations with greater heart ageing (**Figure 5**). In both males and females, the strongest associations were with VAT followed by PAT, though with greater effect sizes in males (VAT: β = 0.115, 95% CI [0.098-0.131], p<0.0001; PAT: β= 0.064 [0.048-0.081], p<0.0001) compared to females (VAT: β = 0.101 [0.086-0.115]; PAT: β = 0.064 [0.049-0.078], both p<0.0001).

**Figure 5.**
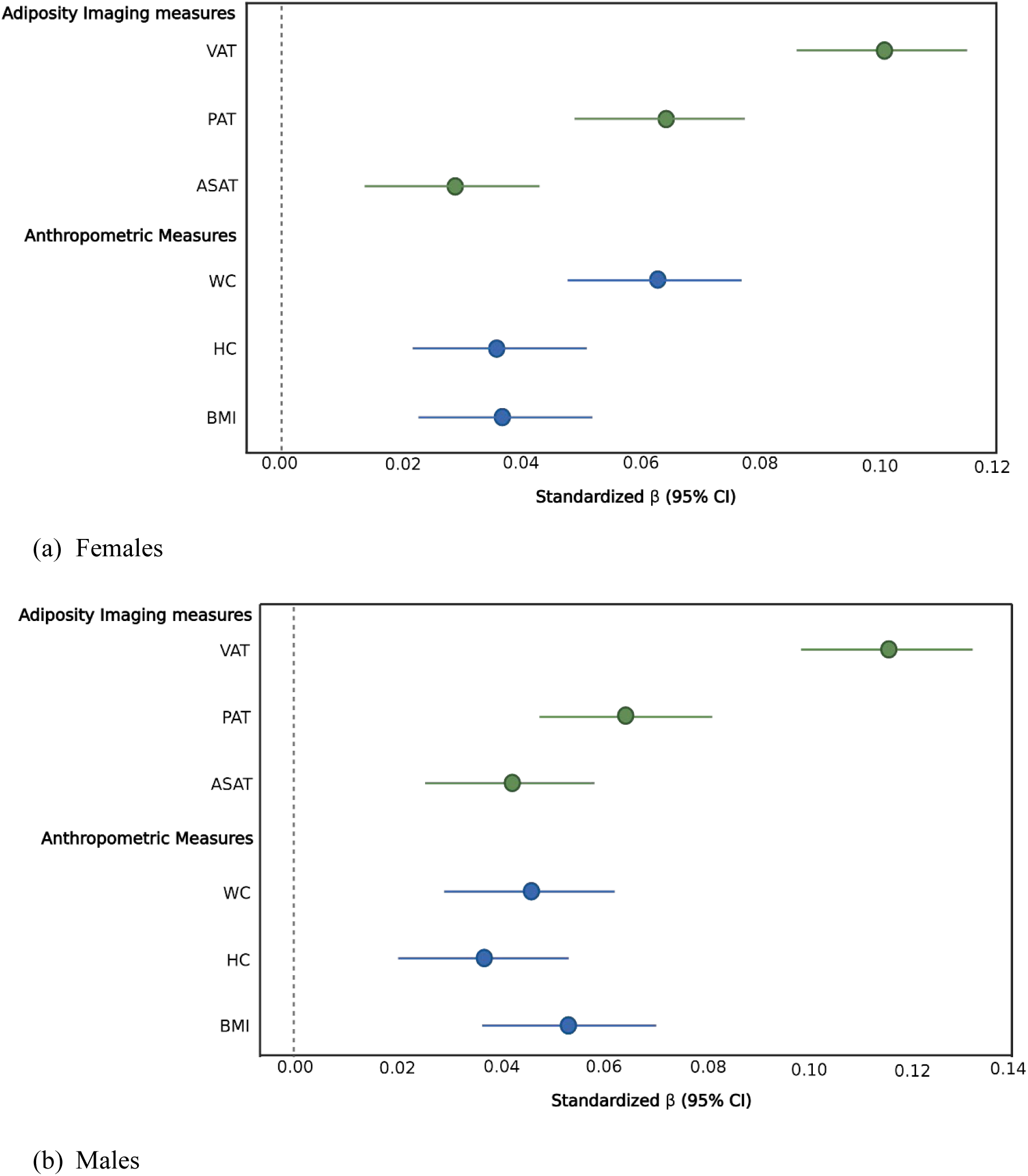
Adiposity-cardiac ageing associations stratified by sex. Forest plots display standardised β coefficients (95% confidence intervals) for (a) females and (b) males. Predictors include WC, VAT, PAT, HC, BMI, and ASAT. Positive effects reflect greater biological cardiac ageing (heart age delta). Dashed vertical line indicates null effect (β = 0). Effects were standardised (z-scores) and adjusted for age, ethnicity, smoking status, and physical activity. Abbreviations: WC, waist circumference; VAT, visceral adipose tissue; PAT, pericardial adipose tissue; HC, Hip circumference; BMI, body mass index; ASAT, abdominal subcutaneous adipose tissue.

Notably, anthropometric measures showed sex-dependent predictive utility. Among females, WC (β = 0.063 [0.048-0.077], p<0.0001) demonstrated stronger associations with cardiac ageing than BMI (β = 0.037 [0.023-0.052], p<0.0001). Conversely, in males, BMI showed relatively stronger predictive value (β = 0.053 [0.037-0.070], p<0.0001) than observed in females.

### Mediation Analysis with incident CVD events

Over an average of 4.71 years of prospective follow-up, there were 811 incident CVD events in males and 550 recorded in females, totalling 1,361 incident CVD cases and 33,135 controls after excluding participants with prevalent CVD. Mediation analyses indicate that heart age plays a significant role in mediating the relationship between adiposity measures and incident CVD events. Notably, VAT demonstrated the strongest mediation effect among the adiposity phenotypes evaluated.

Approximately 13.7% of VAT’s total effect on CVD risk was found to operate through accelerated heart ageing. This was further underscored by the largest indirect effect (1.035) observed among all adiposity measures and the strongest influence of VAT on heart age (β=0.119).

PAT also exhibited an important mediation effect, with 11.9% of its total effect on CVD outcomes explained via heart ageing. This mediation pathway was supported by an indirect effect estimate of 1.022, alongside robust standardised path coefficients from PAT to heart age (β = 0.070) and from heart age to CVD risk (β = 1.354). In contrast, the rest of adiposity measures (BMI, WC, ASAT, and HC), showed comparatively smaller mediation effects. For these, only 6-8% of their total effects on mortality risk were mediated by heart age (**Figure 6**).

**Figure 6.**
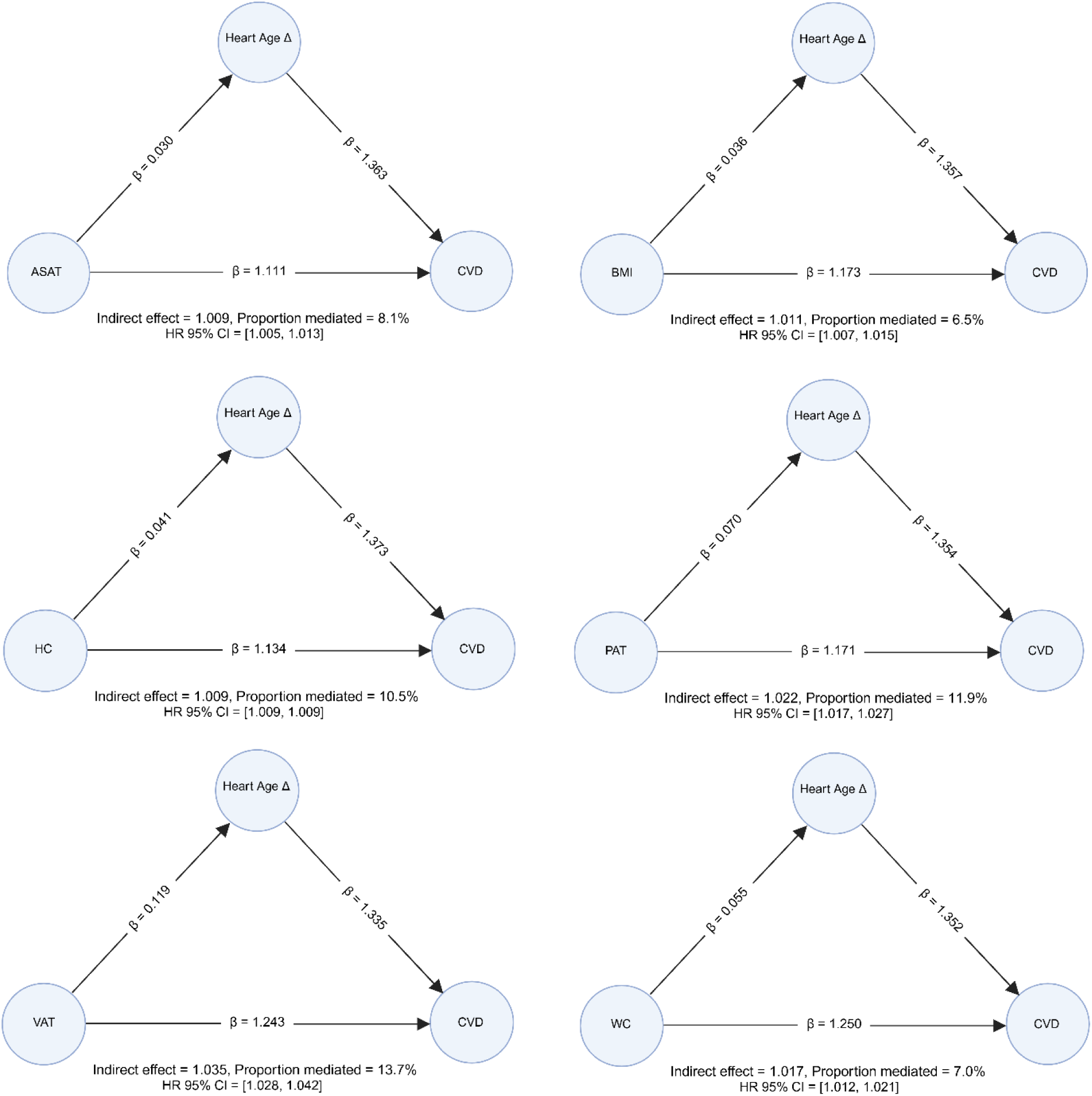
Mediation models for the association between six adiposity-related exposures and CVD, with cardiac age delta as the mediator. Each diagram presents standardised path coefficients (β) for the direct effect of the exposure on CVD, the effect of the exposure on cardiac age delta, and the effect of cardiac age delta on CVD. The indirect effect and corresponding hazard ratio (HR) with 95% confidence interval are reported below each diagram, along with the proportion mediated. β: standardised path coefficient. ASAT, Abdominal Subcutaneous Adipose Tissue; BMI, Body Mass Index; HC, Hip Circumference; PAT, Pericardial Adipose Tissue; VAT, Visceral Adipose Tissue; WC, Waist Circumference; CVD, cardiovascular disease; HR, hazard ratio; CI, confidence interval.

## Discussion

Despite the well-established link between obesity and CVD, traditional anthropometric measures such as BMI often fail to capture the complex relationships underlying adiposity distribution and cardiac health. In this study, we integrated advanced CMR and abdominal MRI biomarkers with ML techniques to characterise compartment-specific obesity phenotypes and evaluate their relationship with biological heart ageing and cardiovascular risk.

Our findings reveal significant sexual dimorphism in the relationship between obesity and cardiac ageing, with males exhibiting consistently stronger detrimental effects covering all adiposity measures. The median heart age delta was significantly higher (indicating greater ageing) in male participants than in female participants (0.15 years vs 0.09, p < 0.001) (**Figure S3**), a pattern that persisted across both clustering-derived phenotypes and conventional BMI categories. Notably, even within the lower-risk cluster, females demonstrated more favourable cardiac ageing profiles than their males’ counterparts (median heart age delta: –0.15 years in females vs. –0.01 years in males), suggesting inherent sex-based differences in physiological resilience to adiposity-driven cardiovascular ageing [15,16]. Our findings are consistent with previous work demonstrating that males have, on average, greater cardiac ageing than females. We further add to the literature, by demonstrating the greater adverse impact of obesity on cardiac ageing in males.

Based on our feature importance analysis, sex-specific patterns in biological heart ageing were evident. In females, LV metrics were the most predictive features. In contrast, right-sided cardiac features, including right ventricular (RV) and right atrial (RA) metrics, ranked among the most important features in males. These findings reflect established understanding of age-related cardiovascular remodelling, and present new information about the differential impact of chamber alterations related to greater biological heart ageing in males and females [14].

Our unsupervised clustering approach revealed two distinct obesity phenotypes, which we labelled as high-risk and low-risk based on their relationship to greater or lower heart ageing. These phenotypes exposed critical limitations of BMI-based classification. Approximately 5% of individuals who were considered to have normal weight by BMI criteria were reclassified into the high-risk cluster due to disproportionately high adiposity levels, particularly VAT, which was over threefold higher in high-risk females compared to their low-risk counterpart. Conversely, 8% of females and 6% of males classified as obese by BMI were assigned to the lower-risk cluster, with heart ageing profiles similar to those of normal-weight individuals. A substantial proportion of overweight individuals were also assigned to the lower-risk group, further underscoring the heterogeneity of obesity and the inadequacy of BMI in capturing cardiovascular risk. Previous work has similar highlighted the added value of image-derived obesity measures, especially VAT and PAT, for more accurate cardiovascular risk stratification beyond BMI [3,6], our findings corroborate these results and extend these observations in the context of heart ageing.

PAT and VAT showed the strongest associations with biological heart ageing in both sexes, with these associations being more pronounced in males. Consistent with our findings, previous studies have demonstrated strong associations between VAT and cardiac ageing in both sexes [6]. Recent work has shown that ectopic and subcutaneous fat depots are strongly associated with accelerated cardiovascular ageing, with some sex-specific effects [16,17]. However, while PAT has also been linked to alterations in left ventricular structure, function, and increased CVD risk [18], its relationship with cardiac ageing has not been previously studied [19,20]. Our results highlight PAT as a distinct and informative imaging-derived feature: its local anatomical proximity to the heart may enable direct paracrine and vasocrine effects, representing a pathway that is not captured by more global measures such as BMI or VAT volume [21]. PAT’s secretory profile is enriched in pro-inflammatory cytokines, adipokines, and profibrotic mediators [22]. These factors can promote myocardial fibrosis, impair diastolic relaxation, and modulate coronary vascular tone, potentially acting independently of systemic adiposity and linking obesity to cardiac ageing and subsequent CVD events [23,24]. The independent association we observed between PAT and accelerated heart ageing suggests that PAT is not merely a passive marker of overall adiposity but an active contributor to the cardiac ageing process. These findings underscore the need for dedicated studies of PAT biology, longitudinal dynamics, and potential interventions to determine whether targeting PAT could slow or reverse age-related cardiac decline.

Previous studies have largely focused on associations between fat depots and age-delta, without examining their downstream impact on clinical outcomes [16]. In contrast, our analysis demonstrates that these fat depots not only relate to heart ageing but also mediate the effect of adiposity on actual CVD events. We found that VAT and PAT were the most important drivers of clinical CVD events mediated through greater cardiac ageing. Our results showed that cardiac ageing mediated 13.7% of the effect of VAT and 11.9% of the effect of PAT on CVD risk. This compares to much lower mediated effects by other metrics which ranged from 6-8%. VAT is thought to exert systemic effects primarily through pathways involving inflammation and insulin resistance [25], whereas PAT has been associated with adverse effects on cardiac mechanics and structure, particularly left ventricular, atrial and right ventricular dysfunction [26,18]. The observation that VAT and PAT contributes more directly to accelerated heart ageing than other fat depots suggests that therapies specifically targeting this fat compartment may mitigate obesity related cardiac decline, even in the absence of substantial weight loss [27,28].

Several limitations warrant consideration. First, the UK Biobank’s demographic bias toward healthier, predominantly European ancestry participants from White ethnicities [29] may limit generalisability, particularly given known demographic and socioeconomic influences on biological ageing [30].

While there was adequate heterogeneity in obesity levels in the samples to conduct reliable analyses and demonstrate associations, there were few people at the extremes of obesity levels, and this may have resulted in underestimation of effect sizes.

This study emphasises the value of integrating advanced imaging biomarkers to disentangle the complex relationships between adiposity and heart ageing. Unsupervised clustering uncovered distinct obesity phenotypes not captured by BMI alone, highlighting the inadequacy of conventional anthropometric measures for cardiometabolic risk stratification. Importantly, heart ageing was shown to partially mediate the effects of both visceral and pericardial fat on cardiovascular risk, positioning heart age delta as a mechanistic link between adiposity and disease. These insights support the need for sex-sensitive, fat-compartment-specific approaches in cardiovascular risk prediction and intervention, potentially informing more precise and effective strategies to combat obesity-related heart disease.

## Methods

### Cohort Description

The UK Biobank is a prospective, population-based cohort study of over 500,000 participants aged 40–69 years at recruitment (2006–2010) [31]. Multi-organ imaging was performed in a large subset.

Longitudinal follow-up is available via linkage to electronic health records, enabling investigation of incident cardiovascular and other chronic diseases. Baseline assessments included detailed questionnaires (socio-demographics, lifestyle behaviours, medical history) and physical measurements [32].

### Obesity measures

#### Anthropometry measurements

BMI (Kg/m^2^) was calculated using height and weight for all participants. Waist circumference (WC) and hip circumference (HC) were obtained at the smallest trunk circumference using a SECA 200-cm tape measure [33]. Participants were categorised into five BMI-based groups: underweight (< 18.5 kg/m^2^), normal weight (18.5 − 24.9 kg/m^2^), overweight (25 − 29.9 kg/m^2^), obesity (30 − 39.9 kg/m^2^), severe obesity (≥ 40 kg/m^2^) [34].

#### Image-Derived Adipose Tissue Quantification

Whole-body MRI was acquired using a Siemens MAGNETOM Aera 1.5-T scanner (Siemens Healthineers, Erlangen, Germany) with a 6-minute dual-echo Dixon Vibe protocol, generating volumetric water/fat-separated datasets (neck to knees) [35]. Body composition analysis, including VAT and ASAT, was performed using AMRA Researcher (AMRA Medical AB, Linköping, Sweden) [36]. Pericardial adipose tissue (PAT) was segmented via an automated, quality-controlled pipeline validated in both the UK Biobank and an external cohort [37].

#### Cardiac magnetic resonance acquisition

CMR was performed using a standardised protocol on 1.5 Tesla Siemens MAGNETOM Aera scanners (Siemens Healthineers, Erlangen, Germany), with consistent equipment and staff training [38]. The imaging protocol included retrospectively gated balanced steady-state free precession (bSSFP) cine sequences acquired in contiguous short-axis stacks (fully covering the left and right ventricles) and long-axis views (horizontal, vertical, left ventricular outflow tract, two-chamber, and four-chamber planes). Typical acquisition parameters were: repetition time/echo time (TR/TE) = 2.6/1.1 ms, flip angle = 80°, parallel imaging (GRAPPA) acceleration factor = 2, and voxel sizes of 1.8 × 1.8 × 8.0 mm (short-axis) or 6.0 mm (long-axis). Temporal resolution was 31–32 ms per phase, interpolated to 50 cardiac phases (∼ 20 ms per phase) for functional analysis. Additional sequences comprised transverse cine imaging of the thoracic aorta and native T1 mapping using a mid-ventricular ShMOLLI sequence [39]. No post-processing filtering was applied beyond manufacturer-provided distortion correction algorithms.

#### Cardiac magnetic resonance measures

We included 56 CMR-derived features to comprehensively characterise cardiac size, function, and tissue properties across all four chambers. These features represent conventional, clinically validated metrics. Parameters of left and right ventricular and atrial size and function included end-diastolic and end-systolic volumes, stroke volumes, ejection fractions, myocardial mass, and global longitudinal strain, offering standardised assessments of chamber dimensions and systolic performance [40].

Tissue characterisation was captured using native myocardial T1 mapping, a non-invasive imaging biomarker of diffuse myocardial fibrosis and other pathological remodelling processes. In addition, vascular compliance was evaluated through ascending aortic distensibility, a marker of central artery stiffness that reflects early vascular deterioration [41].

#### Incident Cardiovascular Disease events

Incident CVD cases were identified through linkage to the Hospital Episode Statistics (HES) database and death registries, coded using the International Classification of Diseases (ICD-9 and ICD-10), as well as through self-reported diagnoses recorded after the baseline imaging assessment (**Table S4**).

#### Covariates and Handling of Missing Data

Primary analyses were adjusted for self-reported ethnicity (white, mixed, South Asian, Chinese, Black, other ethnic group; Data-Field 21000), smoking status (current, never, previous; Data-Field 20116), and frequency of moderate physical activity (number of days per week, ranging from 0 to 7; Data-Field 884). BMI was incorporated through obesity classifications (Data-Field 21001). Unknown values for these covariates were treated as missing (NA). Sex-specific models were used, except in mediation analyses, where sex was included as a covariate. Variables with more than 5% missingness were excluded. Missing data (<5%) for covariates and adiposity measures were handled using k-nearest neighbours’ imputation (k = 10) for continuous variables and mean imputation for discrete variables.

## Statistical methods

### Biological Heart Age Modelling

All analyses were conducted using Python version 3.12.7. For biological heart age estimation, we developed five ML regression models: linear regression, Bayesian ridge, LASSO, XGBoost, and random forest. For model development, we first established a reference standard of healthy cardiovascular ageing by selecting 9,290 females and 5,202 males classified as normal weight (BMI 18.5-24.9 kg/m2) without prevalent CVD. We trained sex-specific models, with each dataset randomly split into training (80%) and test (20%) sets.

For each of the five ML models (linear regression, Bayesian ridge, LASSO, XGBoost, and random forest), optimal hyperparameters were determined through grid search with ten-fold cross-validation on the training set. The model used 56 CMR-derived metrics (ethnicity-adjusted) as predictors and chronological age at imaging as the dependent variable. Performance was evaluated using the validation set to prevent overfitting, using mean absolute error (MAE) and coefficient of determination (R²) as evaluation metrics.

Feature importance was assessed using the built-in XGBoost feature importance plot [42]. In this framework, the F-score quantifies how many times each feature is used to split the data across all trees. Features with higher F-scores contribute more frequently to the model’s decisions and are therefore considered more influential in predicting the outcome.

### Heart Age Estimation and Bias Adjustment

We estimated biological heart age for all participants by calculating the residual between model-predicted heart age and chronological age, termed heart age delta (Δ). This metric provides a quantitative measure of cardiovascular ageing, where: 1) Positive Δ values indicate greater cardiac ageing (heart age > chronological age), 2) Negative Δ values suggest decelerated cardiac ageing (heart age < chronological age) [43].

Consistent with established literature on biological age estimation, we observed systematic regression dilution bias in our heart age predictions [43,44], characterised by three key patterns: (1) underestimation of heart age for older participants, (2) overestimation for younger participants, and (3) the most accurate predictions occurring for individuals near the sample mean age. This bias pattern reflects the well-documented phenomenon in biological age modelling where extreme values tend to be underestimated due to regression toward the mean, while mid-range estimates demonstrate greater accuracy. The observed age-dependent miscalibration necessitated statistical correction to ensure unbiased heart age estimates across the entire age spectrum of our cohort [45,46]. We applied the linear regression-based correction method of [47].

**Stage 1: Bias Characterisation** We computed the linear relationship between initial heart age delta and chronological age in the training set:

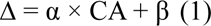

where α (slope) and β (intercept) quantify the systematic prediction bias.

**Stage 2: Age Correction** The trained correction parameters were applied to all predictions:

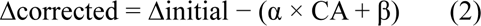

Equivalently, corrected predicted heart age (CPHA) was calculated as:

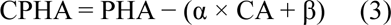

Bias-corrected heart age Δ was used as the primary outcome variable in three complementary analyses. First, we employed unsupervised clustering on adipose tissue imaging features to identify obesity phenotypes. The resulting clusters were then characterised based on heart age Δ to determine whether specific adiposity patterns were associated with accelerated or decelerated cardiac ageing. Second, we examined its associations with conventional anthropometric measures and advanced imaging-derived adiposity metrics. Third, we performed causal inference using mediation analysis to investigate whether heart age Δ mediates the relationship between adiposity metrics and cardiovascular health.

### Obesity clusters

We employed unsupervised k-means clustering (scikit-learn v1.2.0) [48] to identify distinct obesity phenotypes based on body fat distribution patterns. The algorithm partitioned participants using three standardised (z-score) imaging-derived adipose tissue metrics: VAT, ASAT, and PAT. The optimal number of clusters was determined using both the elbow method, which minimises within-cluster variance, and silhouette analysis, where the average score indicated moderate separation.

The two derived clusters were first characterised by their body fat distribution patterns, with median values computed for each imaging-derived adipose tissue metric (VAT, ASAT, PAT) to quantify phenotypic differences. To assess the clinical relevance of these clusters, we evaluated their alignment with conventional BMI-based obesity categories using proportional distributions. Additionally, we compared cardiovascular ageing between clusters by computing the mean heart age delta.

### Associations with Heart Age Delta

To evaluate the relationship between adiposity and cardiovascular ageing, we examined associations between heart age delta bias corrected and comprehensive obesity measures through sex-stratified multivariable linear regression models. Our analysis incorporated both anthropometric assessments (BMI, waist circumference, hip circumference) and advanced imaging-derived adiposity metrics (VAT, ASAT, PAT). All models were adjusted for potential confounders including ethnicity, smoking status, and weekly moderate physical activity. To further characterise these relationships across the adiposity spectrum, we conducted subgroup analyses by obesity category (underweight, normal weight, overweight, obese, and severe obesity) testing for effect modification by adiposity status.

### Mediation analysis

We conducted causal mediation analysis to examine whether heart age delta mediates the association between obesity measures and incident CVD [49]. To increase statistical power, data from both sexes were combined for this analysis, creating a larger and more robust cohort. This approach allowed decomposition of obesity’s total effect on CVD into: (1) a direct effect (independent of heart age delta) and (2) an indirect effect mediated through heart age delta. The analysis incorporated baseline obesity measures (BMI, waist circumference, VAT, ASAT, and PAT) as exposures, bias-corrected heart age delta as the mediator, and time-to-incident CVD (binary) as the outcome, with all models adjusted for age, sex, ethnicity, smoking status, and physical activity. The proportion mediated estimates quantify how much of obesity’s cardiovascular risk operates through accelerated cardiac ageing.

For model specification, we employed linear regression to relate obesity measures to the mediator (heart age delta) and Cox proportional hazards regression for the CVD outcome. Mediation effects were estimated using non-parametric bootstrapping (1,000 iterations) to derive robust 95% confidence intervals, with the proportion mediated calculated as (indirect effect/total effect). Sensitivity analyses addressed potential exposure-mediator interactions and excluded participants with baseline subclinical CVD to minimise reverse causation.

## Data Availability

The scripts for data analysis are publicly available https://anonymous.4open.science/r/Predicting-CVD-using-OCT-images-A586/. Data from UK Biobank is available for approved research.

