## Supplemental Material for "Compartment-specific Fat Distribution Profiles have Distinct Relationships with Cardiovascular Ageing and Future Cardiovascular Events"

**Supplementary Material**

**Online methods**

To ensure reproducibility, we provide the full code for all experiments, including model selection, hyperparameter grid search, and training, in the GitHub repository Github/Cardiac-ageing.

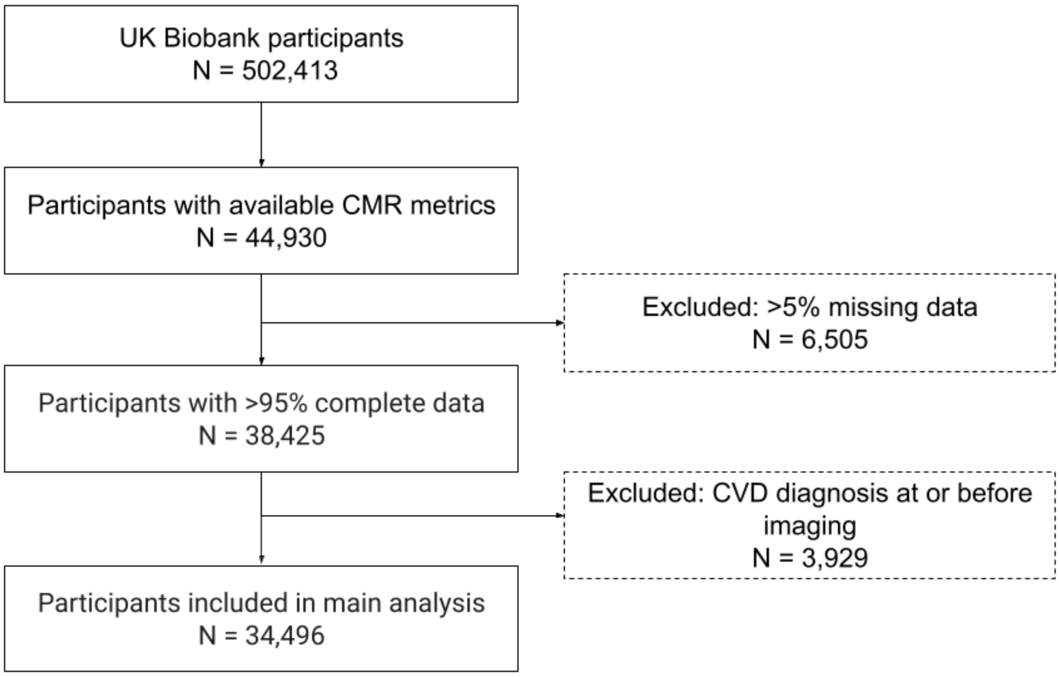


**Figure S1. STROBE flow diagram** illustrating the inclusion and exclusion criteria used to define the study cohort of UK Biobank participants with cardiovascular magnetic resonance (CMR) imaging data.

| Female | | | | | | | | |
|---|---|---|---|---|---|---|---|---|
| | Before correction | | | | After correction | | | |
| Models | MAE | $R^2$ | Corr Delta vs age | Corr pred age vs age | MAE | $R^2$ | Corr Delta vs age | Corr pred age vs age |
| Linear regression | 5.1074 | 0.3044 | -0.7726 | 0.5577 | 3.1252 | 0.7194 | -0.0172 | 0.8820 |
| Bayesian ridge | 5.1053 | 0.3051 | -0.7776 | 0.5576 | 3.0973 | 0.7251 | -0.0170 | 0.8840 |
| LASSO | 5.1001 | 0.3059 | 0.7769 | 0.5877 | 3.0953 | 0.7233 | 0.0364 | 0.8850 |
| XGBoost | 4.9371 | 0.3409 | 0.8039 | 0.5847 | 2.9518 | 0.7647 | 0.01981 | 0.8920 |
| Random Forest | 4.9669 | 0.3382 | 0.7975 | 0.5871 | 3.0115 | 0.7437 | 0.0101 | 0.8890 |
| **Male** | | | | | | | | |
| | Before correction | | | | After correction | | | |
| Models | MAE | $R^2$ | Corr Delta vs age | Corr pred age vs age | MAE | $R^2$ | Corr Delta vs age | Corr pred age vs age |
| Linear regression | 5.5848 | 0.2367 | -0.7966 | 0.5016 | 3.1911 | 0.7221 | 0.0253 | 0.8872 |
| Bayesian ridge | 5.5722 | 0.2437 | -0.8147 | 0.5030 | 3.0626 | 0.7466 | 0.0201 | 0.8950 |
| LASSO | 5.5735 | 0.2394 | -0.8148 | 0.5087 | 3.0953 | 0.7503 | 0.0364 | 0.9017 |
| XGBoost | 5.5578 | 0.2698 | 0.8728 | 0.5205 | 2.6694 | 0.8250 | 0.0511 | 0.9200 |
| Random Forest | 5.5694 | 0.2596 | 0.8640 | 0.5179 | 3.0121 | 0.8130 | 0.036 | 0.9100 |

**Table S1. Performance comparison of heart age delta estimation models before and after bias correction, stratified by sex.** Results show mean absolute error (MAE, years), coefficient of determination (R2), and correlations between predicted/chronological age and heart age delta. Abbreviations: MAE, mean absolute error; Corr, Pearson correlation coefficient.

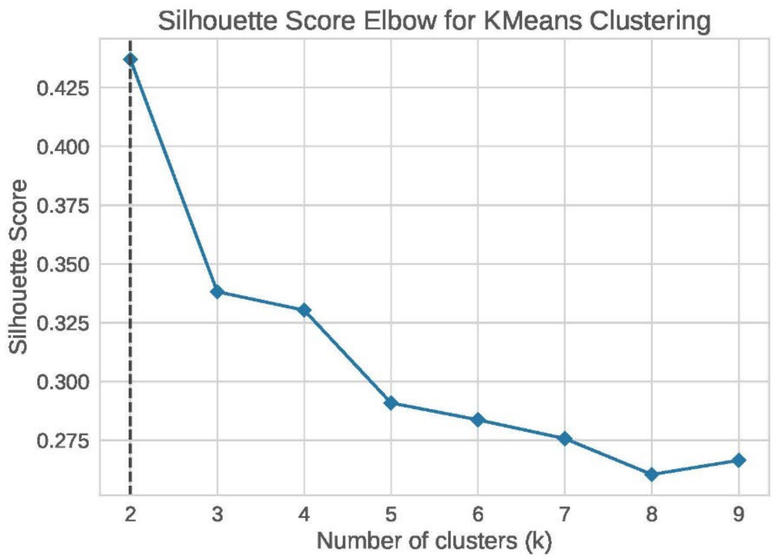


(a) Females

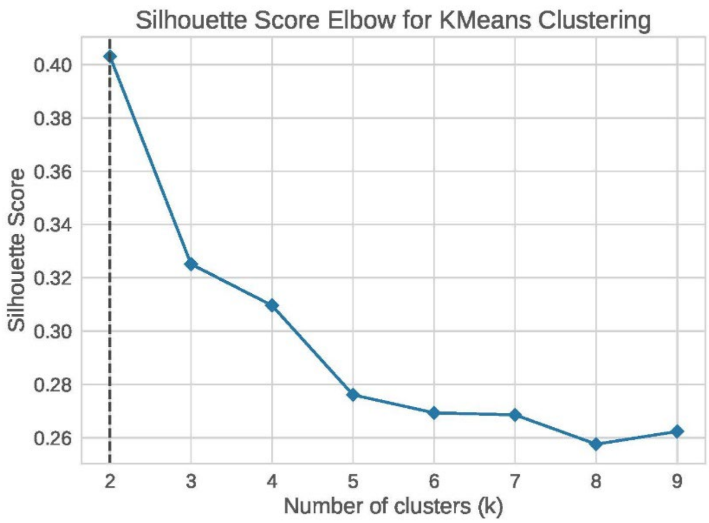


(b) Males

**Figure S2. Adiposity-cardiac ageing associations stratified by sex.** Forest plots display standardised β coefficients (95% confidence intervals) for (a) females and (b) males. Predictors include WC, VAT, PAT, HC, BMI, and ASAT. Positive effects reflect greater biological cardiac ageing (heart age delta). Dashed vertical line indicates null effect (β = 0). Effects were standardised (z-scores) and adjusted for age, ethnicity, smoking status, and physical activity. Abbreviations: WC, waist circumference; VAT, visceral adipose tissue; PAT, pericardial adipose tissue; HC, Hip circumference; BMI, body mass index; ASAT, abdominal subcutaneous adipose tissue.

| Female | | | | |
|---|---|---|---|---|
| Cluster 0 | | | | |
| | VAT | ASAT | PAT | N |
| Obesity Group | | | | |
| Underweight | - | - | - | 0 |
| Normal weight | 3.28 (0.78) | 7.32 (1.25) | 29.08 (6.95) | 459 |
| Overweight | 3.76 (0.94) | 9.57 (1.61) | 25.17 (7.73) | 2919 |
| Obesity | 4.57 (1.28) | 12.76 (2.36) | 26.22 (9.73) | 2713 |
| Severe obesity | 6.001 (1.55) | 18.54 (2.50) | 30.59 (11.27) | 226 |
| Cluster 1 | | | | |
| | VAT | ASAT | PAT | N |
| Obesity Group | | | | |
| Underweight | 0.71 (0.32) | 2.50 (0.79) | 11.71 (4.90) | 265 |
| Normal weight | 1.60 (0.69) | 5.44 (1.56) | 15.16 (5.46) | 8652 |
| Overweight | 2.27 (0.67) | 8.001 (1.40) | 15.47 (4.69) | 3520 |
| Obesity | 2.49 (0.61) | 9.91 (1.40) | 14.20 (3.80) | 224 |
| Severe obesity | - | - | - | 0 |
| **Male** | | | | |
| Cluster 0 | | | | |
| | VAT | ASAT | PAT | N |
| Obesity Group | | | | |
| Underweight | - | - | - | 0 |
| Normal weight | 5.55 (1.10) | 5.04 (0.94) | 44.50 (10.24) | 259 |
| Overweight | 6.32 (1.29) | 6.52 (1.36) | 40.11 (11.91) | 3386 |
| Obesity | 7.64 (1.76) | 9.07 (2.35) | 40.15 (14.53) | 2394 |
| Severe obesity | 10.30 (2.47) | 16.14 (3.28) | 44.28 (14.08) | 100 |
| Cluster 1 | | | | |
| | VAT | ASAT | PAT | N |
| Obesity Group | | | | |
| Underweight | 1.24 (0.56) | 1.92 (0.62) | 15.00 (6.83) | 36 |
| Normal weight | 2.79 (1.19) | 3.80 (1.07) | 20.87 (8.59) | 5080 |
| Overweight | 4.17 (1.13) | 5.30 (1.14) | 23.65 (7.74) | 4088 |
| Obesity | 4.86 (1.03) | 6.51 (1.21) | 21.32 (6.04) | 175 |
| Severe obesity | - | - | - | 0 |

**Table S2. Sex-stratified adiposity measures by phenotypic cluster and BMI subgroup.** Values represent mean (standard deviation) for visceral adipose tissue (VAT, mL), abdominal subcutaneous adipose tissue (ASAT, L), and pericardial adipose tissue (PAT, $cm^2$) across phenotypic clusters (Cluster 0 = high-risk phenotype; Cluster 1 = protected phenotype) within each BMI subgroup. Data are presented separately for females and males.

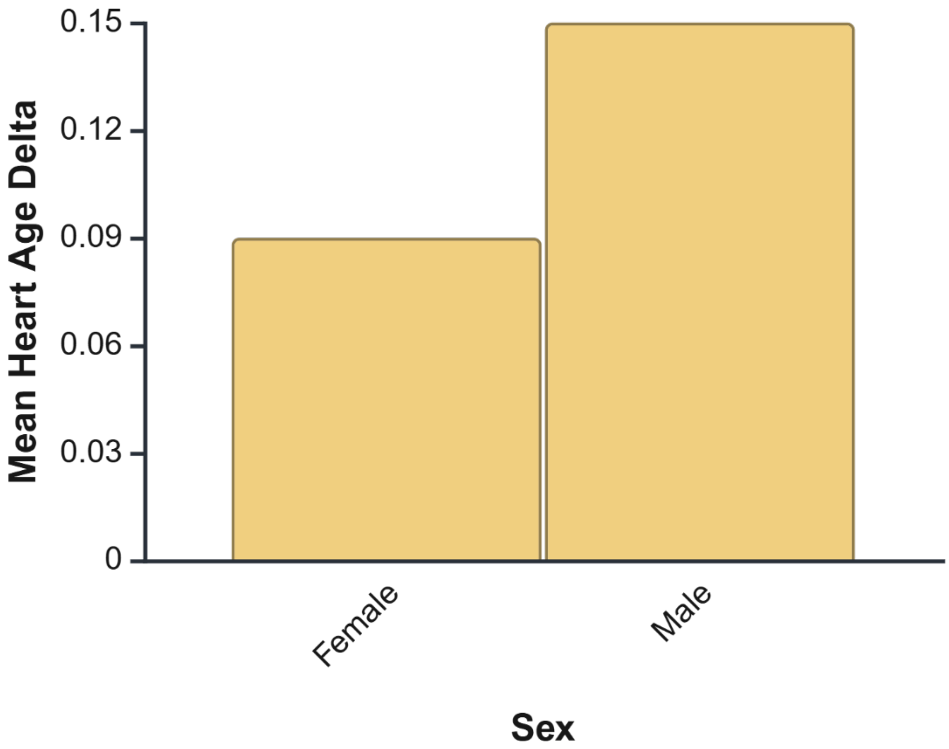


**Figure S3. Mean heart age delta stratified by sex.** Positive values indicate higher cardiac ageing (heart age > chronological age), while negative values indicate decelerated ageing.

| Exposure | Total Effect | Direct Effect | Indirect Effect | $\beta(X \rightarrow M)$ | $\beta(M \rightarrow Y)$ | Proportion Mediated |
|---|---|---|---|---|---|---|
| ASAT | 1.121 | 1.111 | 1.009 | 0.030 | 1.363 | 0.081 |
| BMI | 1.186 | 1.173 | 1.011 | 0.036 | 1.357 | 0.065 |
| HC | 1.120 | 1.134 | 1.010 | 0.041 | 1.353 | 0.105 |
| PAT | 1.196 | 1.171 | 1.022 | 0.070 | 1.354 | 0.119 |
| VAT | 1.287 | 1.243 | 1.035 | 0.119 | 1.335 | 0.137 |
| WC | 1.171 | 1.150 | 1.017 | 0.055 | 1.352 | 0.070 |

**Table S3. Mediation analysis evaluating the role of heart age as a mediator in the association between various adiposity measures and CVD.** Estimates represent standardised effects from mediation models evaluating heart age as a mediator (M) of the relationship between each adiposity measure (X) and mortality risk (Y). The path from X to M was estimated using linear regression adjusted for covariates (ethnicity, number of days for physical activity, sex, smoking status), while the path from M to Y and the direct effect of X on Y were estimated using a Cox proportional hazards model. Proportion mediated is calculated as the ratio of the indirect effect to the total effect. Abbreviations: WC, waist circumference; VAT, visceral adipose tissue; PAT, pericardial adipose tissue; HC, Hip circumference; BMI, body mass index; ASAT, abdominal subcutaneous adipose tissue.

| | UKB Field ID / Code | Description |
|---|---|---|
| **Stroke** | | |
| Self-report | 20002 | Stroke, Ischaemic stroke, Brain hemorrhage |
| ICD9 | 431<br>432 | Intracerebral hemorrhage<br>Other and unspecified intracranial hemorrhage |
| ICD10 | I64<br>I63<br>I61<br>I62 | Stroke, not specified as hemorrhage or infarction<br>Cerebral infarction<br>Intracerebral hemorrhage<br>Other nontraumatic intracranial hemorrhage |
| Algorithm | 42006<br>42008<br>42010 | Date of stroke<br>Date of ischaemic stroke<br>Date of intracerebral hemorrhage |
| **Cardiac arrhythmia** | | |
| Self-report | 20002 | Sick sinus syndrome, SVT / supraventricular tachycardia, Atrial flutter, Heart arrhythmia, Irregular heart beat, Atrial fibrillation |
| ICD10 | I44.1<br>I44.2<br>I45.3<br>I45.6<br>I46.0<br>I46.1<br>I46.9<br>I47.0<br>I47.1<br>I47.2<br>I47.9<br>I48.0<br>I48.1<br>I48.2<br>I48.3<br>I48.4<br>I48.9<br>I49.0<br>I49.5 | Atrioventricular block, second degree<br>Atrioventricular block, complete<br>Trifascicular block<br>Preexcitation syndrome<br>Cardiac arrest with successful resuscitation<br>Sudden cardiac death, so described<br>Cardiac arrest, unspecified<br>Re-entry ventricular arrhythmia<br>Supraventricular tachycardia<br>Ventricular tachycardia<br>Paroxysmal tachycardia, unspecified<br>Paroxysmal atrial fibrillation<br>Persistent atrial fibrillation<br>Chronic atrial fibrillation<br>Typical atrial flutter<br>Atypical atrial flutter<br>Atrial fibrillation and atrial flutter, unspecified<br>Ventricular fibrillation and flutter<br>Sick sinus syndrome |
| **Heart failure** | | |
| Self-report | 20002 | Heart failure/pulmonary oedema |
| ICD10 | I50.0 | Congestive heart failure |
| | I50.1 | Left ventricular failure |
| | I50.9 | Heart failure, unspecified |
| **Ischaemic heart disease** | | |
| Self-report | 20002 | Angina, Heart attack/myocardial infarction |
| ICD9 | 410 | Acute myocardial infarction |
| | 411 | Other acute and subacute forms of ischaemic heart disease |
| | 412 | Old myocardial infarction |
| ICD10 | I20 | Angina pectoris |
| | I21 | Acute myocardial infarction |
| | I22 | Subsequent myocardial infarction |
| | I23 | Certain current complications following acute myocardial infarction |
| | I24 | Other acute ischaemic heart diseases |
| | I25 | Chronic ischaemic heart disease |

| Source | UKB Field ID / Code | Description |
|---|---|---|
| **Non-ischaemic cardiomyopathies** | | |
| Self-report | 20002 | Cardiomyopathy, Hypertrophic cardiomyopathy (HCM / HOCM) |
| ICD10 | I42 | Cardiomyopathy |
| | I43 | Cardiomyopathy in diseases classified elsewhere |
| | I11 | Hypertensive heart disease |
| | I13 | Hypertensive heart and renal disease |
| **Valvular heart disease** | | |
| Self-report | 20002 | Mitral stenosis, mitral valve disease, heart valve problem/heart murmur, mitral regurgitation / incompetence, aortic valve disease, aortic stenosis, aortic regurgitation / incompetence |
| ICD10 | I34 | Mitral (valve) insufficiency |
| | I34.2 | Non-rheumatic mitral (valve) stenosis |
| | I34.8 | Other nonrheumatic mitral valve disorders |
| | I34.9 | Non-rheumatic mitral valve disorder, unspecified |
| | I35 | Non-rheumatic aortic valve disorders |
| | I36 | Non-rheumatic tricuspid valve disorders |
| | I37 | Pulmonary valve disorders |
| | I38 | Endocarditis, valve unspecified |
| | I39.0 | Mitral valve disorders in diseases classified elsewhere |
| | I39.1 | Aortic valve disorders in diseases classified elsewhere |
| | I39.3 | Pulmonary valve disorders in diseases classified elsewhere |
| | I39.4 | Multiple valve disorders in diseases classified elsewhere |
| | I39.8 | Endocarditis, valve unspecified, in diseases classified elsewhere |
| | I05 | Rheumatic mitral valve diseases |
| | I06 | Rheumatic aortic valve diseases |
| | I07 | Rheumatic tricuspid valve diseases |
| | I08 | Multiple valve diseases |

**Table S4. UK Biobank Cardiovascular Diseases definitions.**
